# From Past to Present: Pompe Disease, Pseudodeficiency Alleles, and Diagnostic Challenges

**DOI:** 10.1101/2024.10.03.24314698

**Authors:** Florencia Giliberto, Paula Inés Buonfiglio, Gabriel Capellino, Carmen Llames Massini, Viviana Dalamón, Leonela Luce, Micaela Carcione

## Abstract

Pompe disease is an autosomal recessive disorder caused by *GAA* variants leading to acid alpha-glucosidase deficiency. Diagnosis is challenging due to the variable phenotypic presentation and overlap with other conditions. Traditionally, diagnosis relies on measuring enzyme activity, but next-generation sequencing (NGS) advancements have improved accuracy. However, interpreting variants is complex, especially because pseudodeficiency alleles mimic disease-causing variants. We present two patients harboring the pseudodeficiency allele NM_000152.5(*GAA*):c.271G>A, p.Asp91Asn, which is confusing due to inaccurate reports and results related to enzymatic activity. The first case was a recently published controversial case of a 700-year-old mummy in which the authors classified the variant as pathogenic. The second patient had symptoms compatible with late-onset Pompe disease and was homozygous for the variant. We aimed to determine the correct variant classification using *GAA*:c.271G>A as a model and to achieve a genetic diagnosis of the second patient. This variant was analyzed following international guidelines (ACMG-AMP) and reviewed with the Lysosomal Diseases Variant Curation Expert Panel. The second patient underwent NGS. We demonstrated that *GAA*:c.271G>A meets the criterion of being classified as benign for Pompe. Additionally, the second patient carried a heterozygous pathogenic *PABPN1* variant associated with oculopharyngeal muscular dystrophy, which better explained the clinical features. This underscores the importance of expanding the genetic analysis in the presence of pseudodeficiency alleles that can mask the true cause of the disease and highlights the fact that an accurate diagnosis should adhere to guidelines on variant curation to reduce the risk of misdiagnosis, which could result in inadequate care and risky medical decisions.

**Graphical abstract:** 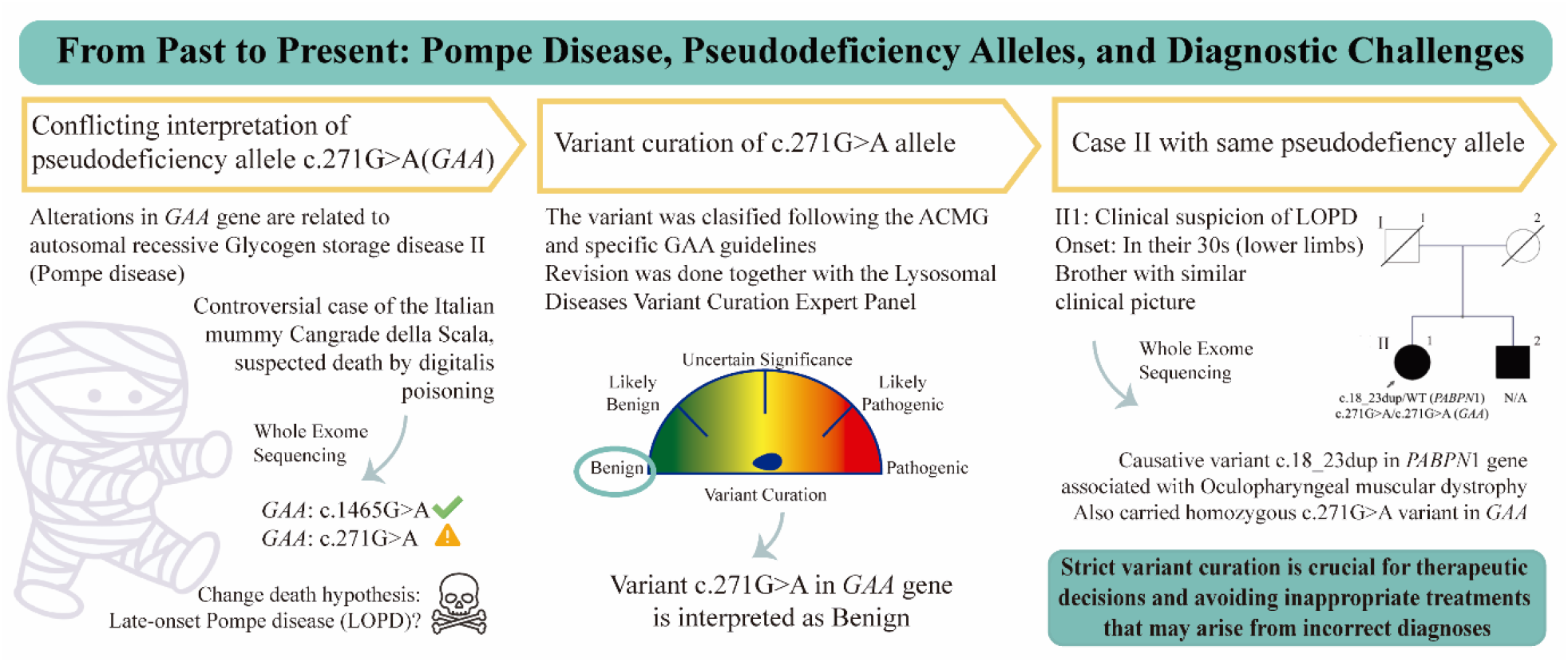

## Introduction

Pompe disease, also named glycogen storage disease type II or acid maltase deficiency, is a rare genetic disorder with an autosomal recessive inheritance pattern. Globally, the prevalence of this disease is estimated to be between 5,000 and 10,000 people, with an incidence of 1 in 57,000 people for the late-onset form [1]. Pompe disease is caused by alterations in the *GAA* gene, leading to a deficiency of acid alpha-glucosidase, a crucial lysosomal enzyme for degrading glycogen [2–4]. Its deficiency causes glycogen accumulation in lysosomes, leading to progressive cellular dysfunction and cell death. It primarily affects neurons and skeletal, smooth and cardiac muscles, resulting in muscle function impairment and progressive and irreversible deterioration [5].

Pompe disease exhibits significant clinical heterogeneity, variation is based on the age of onset and symptoms progression. Residual acid alpha-glucosidase activity determines the onset of the disease. Infantile-onset Pompe disease (IOPD) is the most severe form of the disease and occurs when the enzyme activity is less than 1%, conversely, late-onset Pompe disease (LOPD) occurs when the residual enzyme activity ranges between 2% and 30% [4]. Symptoms of IOPD include progressive muscle weakness, hypotonia since birth, severe respiratory problems, hypertrophic cardiomyopathy, and hepatomegaly. Patients also often experience delays in motor development, such as difficulties in sitting, crawling, or walking [2,5]. However, symptoms in LOPD patients can have a juvenile or adult onset, and patients tend to develop symptoms more slowly than IOPD patients. Patients may present progressive muscle weakness, respiratory problems that worsen in advanced stages, muscle pain, fatigue, and difficulty in walking. Muscle atrophy and proximal weakness are also common features of this disease [1].

The type of molecular alteration and its location in the *GAA* gene often predict the clinical phenotype of Pompe disease [6]. To date, more than 300 alterations in *GAA* have been described [2–4]. Most (96%) of them are single nucleotide variants (SNVs), while only 4% are copy number variants (CNVs). Therefore, sequencing is the ideal strategy for executing the *GAA* genetic diagnostic algorithm [7,8]. The measurement of acid alpha-glucosidase activity in dried blood spots was traditionally the primary method for diagnosing Pompe disease. In recent years, the advances and popularization of NGS technologies have led to a paradigm shift toward genetic/molecular diagnosis. To reduce arbitrariness in the classification of identified sequence variants, the American College of Medical Genetics and Genomics (ACMG) and the Association for Molecular Pathology (AMP) have developed international consensus guidelines to estimate a score based on collected evidence that allows variant classification into one of five possible categories: pathogenic (≥10), likely pathogenic (6 to 9), uncertain significance (0 to 5), likely benign (−1 to -6), or benign (≤-7) [9]. Current recommendations indicate that the diagnosis of Pompe disease should be established when both a deficit of acid alpha-glucosidase enzyme and two pathogenic *trans* variants in the *GAA* gene are detected [10]. Accurate and early diagnosis of Pompe disease is even more relevant now that an enzyme replacement therapy for treating these patients has been approved [11,12].

It should be noted that diagnosing Pompe disease can be challenging. Clinically speaking, the variable presentation of the disease and the symptoms overlap with other conditions and often delay diagnosis. Additionally, the interpretation of *GAA* variants can be complex, especially in the presence of pseudodeficiency alleles, which can mimic disease-causing variants. The main difficulty lies in the fact that pseudodeficiency alleles exhibit low acid alpha-glucosidase enzyme activity in *in vitro* tests, which can be indistinguishable from the pathological deficiency observed in these tests [13]. Therefore, additional biochemical analyses are required to distinguish between true enzyme deficiency leading to Pompe disease and pseudodeficiency alleles that have no clinical impact [10]. To facilitate variant interpretation, the ClinGen Lysosomal Diseases Variant Curation Expert Panel recently adapted the ACMG-AMP guidelines for the classification of molecular alterations involved in metabolic storage diseases and the development of specific criteria for the analysis of *GAA* variants [14].

Here, we present two cases in which the pseudodeficiency allele NM_000152.5(*GAA*):c.271G>A, p.Asp91Asn, also known as *GAA2*, was identified. The first is a controversial case of a 700-year-old mummy published in 2021, where the authors classified the change as pathogenic and as an LOPD case, confusing the interpretation of the variant within the scientific community [15]. The second patient referred to our laboratory was clinically suspected of presenting LOPD. We demonstrated the importance of adhering to international guidelines for accurate classification of variants that follow Mendelian inheritance using this pseudodeficiency allele as a model. Finally, our goal was to achieve a definitive and accurate diagnosis of the second patient, underscoring the critical role of precise genetic diagnosis in guiding appropriate clinical management.

## Materials and Methods

### Patient

The patient, hereafter called “Case 2”, was referred by a neurologist to the Dystrophinopathies Laboratory (INIGEM, UBA-CONICET-Hospital de Clínicas “José de San Martín”) with a clinical suspicion of Pompe disease. Muscle strength was evaluated manually using the Medical Research Council (MRC) scale [16]. Alpha-glucosidase activity was measured from dried blood spots, with reference values between 2.10 and 29.00 μmol/L/h. The protocol was approved by the Institutional Review Board. Informed consent was obtained from the study subjects prior to the molecular studies.

### Samples and NGS Sequencing

Genomic DNA (gDNA) was extracted from peripheral blood lymphocytes of patient 2 following the cetyltrimethylammonium bromide (CTAB) protocol [17]. Then, whole exome sequencing (WES) was performed using the Illumina NovaSeq Sequencing Platform and the Agilent SureSelect V6-post as a capture kit (coverage>20X: 91.7%; Macrogen Service, Korea). Aligned reads were compared to the human reference genome assembly GRCh38. WES results were filtered by an *in silico* panel that included the approximately 700 genes described in the work titled “The 2024 version of the gene table of neuromuscular disorders (nuclear genome)” [18]. Variant filtering and analysis consisted of evaluating several parameters, such as inheritance patterns, variant location, variant type (e.g., missense variants, splice site alterations, and coding in-frame and nonframeshift indels), and variant frequency in population databases such as gnomAD [https://gnomad.broadinstitute.org/]. Published reports of pathogenicity and data from resources such as ClinVar [https://www.ncbi.nlm.nih.gov/clinvar/] were also integrated. Pathogenicity predictions were performed using REVEL [https://sites.google.com/site/revelgenomics. The gathered data were synthesized, and variant classifications were determined using tools such as Varsome [https://varsome.com/] and the Franklin Platform [https://franklin2.genoox.com/]. All the implemented databases, tools and platforms were last accessed on 26.08.2024. [9]

### Classification of variants

Variants were classified following the international ACMG-AMP guidelines [9]. The variant NM_000152.5*(GAA):*c.271G>A, p.Asp91Asn, was assessed by applying *the* latest specific guidelines from the Lysosomal Diseases Expert Panel (ClinGen LSD VCEP) [https://clinicalgenome.org/affiliation/50009/] [14].

## Results

### Case 1: Clinical characteristics

The proband is the Cangrande della Scala (1291–1329 CE), a prominent figure from medieval Italy known for his military and political accomplishments as the Lord of Verona. Due to the destruction of the Scaliger family archives, the little we know about his health relies on limited, ambiguous and, often, biased historical accounts. Contemporary sources indicate that Cangrande suffered from several bouts of illness throughout his life. At the age of 23, he presented a significant ailment that temporarily hindered his mobility. Later, at the age of 34, he suffered from a severe and prolonged illness that was nearly fatal. Last, at the age of 38 years, he died from an illness vaguely described as *fluxum*, often interpreted as vomiting or diarrhea. The accounts report that his last three days were marked by fever and gastrointestinal symptoms. In 2004, an autopsy and a toxicology analysis performed on Cangrande’s mummified remains detected toxic doses of digitalis in his preserved organs, which could be interpreted as poisoning or as a remedy for a cardiac condition. Overall, the above mentioned accounts are nonspecific and could be due to various conditions, ranging from battle injuries to infectious or chronic diseases.

### Case 2: Clinical characteristics

The proband (II.1) had an onset in their 30s, with proximal weakness, predominantly in the lower limbs. Physical examination revealed 4/5 strength in the flexors and extensors of the neck, deltoids, biceps, triceps, psoas, quadriceps, gluteal muscles, hamstrings, and abdominal muscles, with an inability to stand up. Reflexes were preserved. The patient reported swallowing difficulties. Muscle MRI revealed moderate atrophy with fatty infiltration in the gluteal and hamstring muscles. During their last evaluation, in their 70s, the patient exhibited 4/5 weakness in the deltoids, biceps, eyelids, quadriceps, psoas, and gluteal muscles. The alpha-glucosidase activity was 1.68 μmol/L/h, which was slightly lower than the reference value.

The patient’s brother (II.2) exhibited a similar clinical picture, but he declined to participate in the genetic testing (Figure 1A). The proband reported that her deceased parents showed no visible signs of muscle weakness compatible with LOPD.

**Figure 1.**
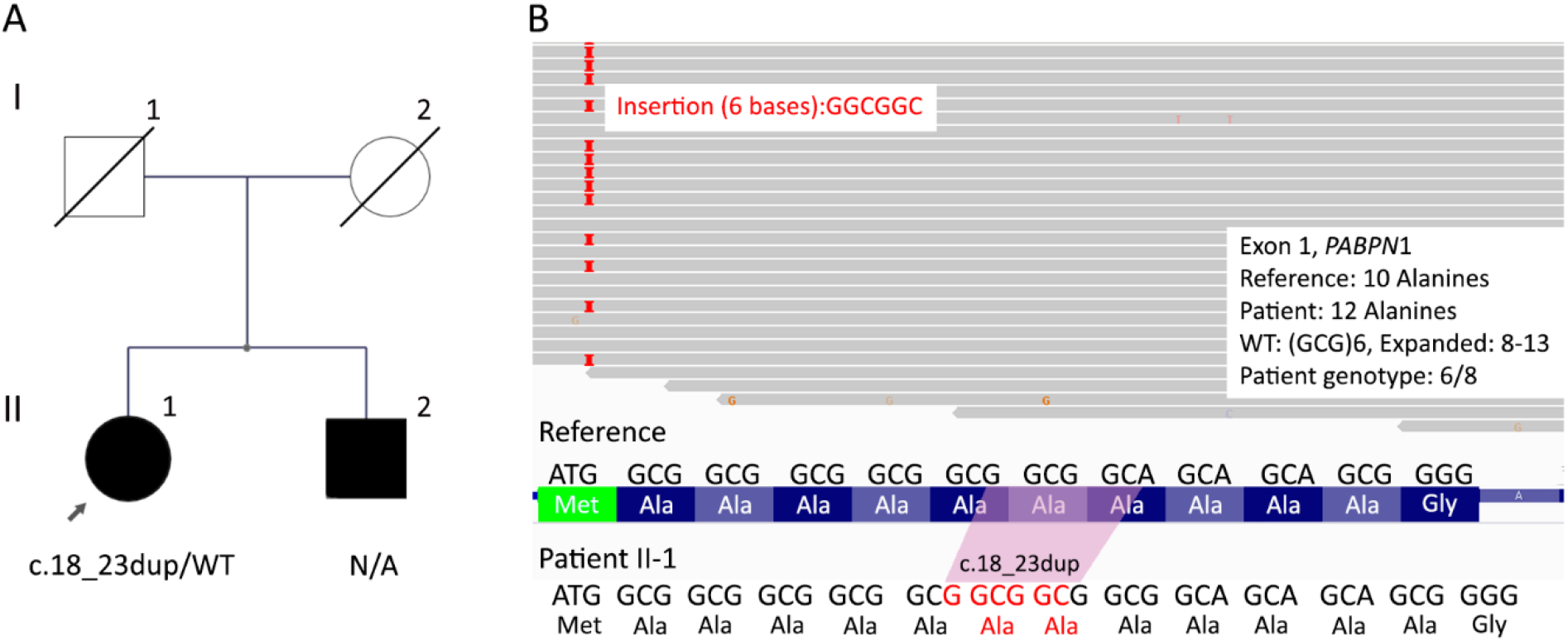
Case 2 presentation. A: Pedigree of the affected family. The proband is indicated by an arrow. N/A: Not available. WT: wild-type allele. B: Visualization of the proband’s sequencing reads via IGV, demonstrating the expansion of the GCG triplet in exon 1 of the *PABPN*1 gene, resulting in two additional alanines in the protein.

### Classification of the NM_000152.5(GAA):c.271G>A, p.Asp91Asn variant

The *GAA2* variant was found in a heterozygous state together with the pathogenic NM_000152.5(GAA):c.1465G>A in the mummy from case 1 and in a homozygous state in case 2. Therefore, to reclassify the variant and provide genetic diagnosis, evidence was gathered from various literature reviews and databases and rigorously analyzed. A summary of the applied criteria is provided in Table 1. Notably, there is evidence indicating that the *GAA2* variant affects splicing and *in vitro* enzymatic activity; however, this effect was proven to be compensated by an increase in substrate concentration [13,19,20]. Therefore, neither the PS3 nor the BS3 criteria were applied. Additionally, BA1 was implemented given the variant’s high allele frequency in general population databases, a criterion that could not be excluded owing to the lack of strong and solid evidence supporting its pathogenicity. Overall, the *GAA2* variant meets the criterion of being classified as benign for Pompe disease. Moreover, the correlation between the presence of *GAA2* and decreased alpha-glucosidase activity indicated that the variant was a pseudodeficiency allele.

**Table 1:** Summary of the rules applied for the interpretation of the variant NM_000152.5(*GAA*):c.271G>A (p.Asp91Asn)

| Applied Rules | Explanation | Reference/Evidence |
| --- | --- | --- |
| BA1<br>(Stand alone) | Allele frequency is 3.29% (4183/127102, 70 homozygous in non-Finnish European population) in GnomAD v2.1.1 | [gnomad.broadinstitute.org] |
| BP2<br>(Supporting) | Observed in Cis with a pathogenic variant in patients with POMPE. | PMID: 16838077, 33073003, 27189384(28–30) |
| BP4<br>(Supporting) | REVEL score of 0.396, which is below the threshold of 0.5 for the pathogenicity effect. The SpliceAI splicing predictor predicts that the variant has no impact on splicing. | Revel and SpliceAI predictor scores were determined with the Variant Effect Predictor tool [https://www.ensembl.org/Tools/VEP] |
| BS2 (Strong) | Observed in a healthy adult individual for a recessive (homozygous), dominant (heterozygous), or X-linked (hemizygous) disorder, with full penetrance expected at a young age. | [gnomad.broadinstitute.org] |

### Classification of the NM_004643.4 (PABPN1):c.18_23dup variant

To provide a genetic diagnosis for patient 2 and due to the reclassification of the *GAA2* variant as benign, the WES analysis was expanded to other genes with clinical features overlapping with LOPD. This screening revealed a heterozygous 6-nucleotide duplication in *PABPN1* [NM_004643.4 (*PABPN1*):c.18_23dup]. This gene is associated with autosomal dominant oculopharyngeal muscular dystrophy (OPMD; OMIM #602279), a disease caused by the abnormal expansion of GCN trinucleotide repeats. The detected duplication resulted in an expansion of the GCG triplet, leading to the addition of two alanine residues in the polyalanine tract region (Figure 1). Evidence was gathered to classify the variant, and a score of 10 points was achieved, reaching the pathogenic classification. A summary of the applied criteria is provided in Table 2. Therefore, the proband was diagnosed with OPMD instead of the suspected late-onset Pompe disease.

**Table 2:** Summary of the rules applied for the interpretation of the variant *PABPN1*(NM_004643.4):c.18_23dup (p.Ala10_Ala11dup)

| Applied Rules | Explanation | Reference/Evidence |
| --- | --- | --- |
| PM1 (Moderate) | Located in a critical and well-established mutational hot spot and/or functional domain (e.g., active site of an enzyme) without benign variation. | 11 reported pathogenic or probably pathogenic variants were found in a 25 bp region surrounding this variant in exon 1 without any benign variants. |
| PS1 (Strong) | Same amino acid change as a variant previously reported as pathogenic | High-certainty ClinVar pathogenic variants. Variant ID: 2437738; Chr14:23321487:G>GGCGGA |
| PS4 (Moderate) | The prevalence of the variant in affected individuals is significantly higher compared to that in controls. | Reported in ClinVar in cases of affected SCV000330133: "The c.18_23dup pathogenic variant in the *PABPN1* gene has been reported previously in individuals with oculopharyngeal muscular dystrophy in four families (Brais et al. 1998)" (26)<br>SCV001248587: "Reported in affected case by submitting lab" |
| PM2 (Supporting) | Extremely low frequency in gnomAd population databases | [gnomad.broadinstitute.org] |
| PP4 (Supporting) | Patient's phenotype or family history is highly specific for a disease with a single genetic etiology. | The c.18_23dup pathogenic variant in the *PABPN1* gene has been reported previously in individuals with oculopharyngeal muscular dystrophy in four families (26) |

## Discussion

For several years, the interpretation of the *NM_000152*.*5(GAA):c*.*271G>A pseudodeficiency allele has led to controversy among the* scientific community due to its inaccurate *in vitro* enzymatic activity [13,19–21]. Iadarola *et al*. (2021) further contributed to this confusion by classifying the variant as pathogenic in a renowned journal in 2021 [15]. Furthermore, the authors concluded that Cangrande della Scala suffered from LOPD; thus, he was the first known case of this disease. These authors based their conclusions on the interpretation of insufficient, imprecise and biased clinical data obtained from limited reliable sources over 7 centuries of age, together with the finding of two variants in the *GAA* gene (the pseudodeficiency allele c.271G>A and a pathogenic variant c.1465G>A) in the mummified sample. According to the current diagnostic guidelines for Pompe disease, which requires both a decreased GAA enzymatic activity and two pathogenic GAA *in trans* variants, there are several reasons why it is incorrect to assert that Cangrande had LOPD [10,22,23]. First, due to the nature of the sample, it was not possible to measure the alpha-glucosidase activity. Second, the authors did not prove with 100% certainty that the *GAA* changes were in trans. Third and most importantly, based on the evidence available to date Cangrande, the mummy, carried a single heterozygous pathogenic *GAA* variant, and the NM_000152.5(*GAA*):c.271G>A, p.Asp91Asn, which is interpreted as benign for Pompe disease. Therefore, based on the available historical data and autopsy findings, there is no clear or compelling evidence sufficient to determine that Cangrande della Scala suffered from Pompe disease or any other specific lysosomal storage disorder. The existing medical records and interpretations remain vague and insufficient to draw definitive conclusions about underlying health issues. The most important outcome from the reanalysis of this case is that, in collaboration with the ClinGen Lysosomal Storage Disorders Expert Panel, the variant was successfully reclassified as benign in ClinVar [https://www.ncbi.nlm.nih.gov/clinvar/variation/4020/].

In the second case, the patient, with clinical suspicion of LOPD, was genetically diagnosed with OPMD owing to a pathogenic heterozygous variant in polyadenylate-binding protein nuclear 1 (PABPN1). The protein encoded by *PABPN1* plays a crucial role in adding poly(A) tails to the 3’ ends of pre-messenger RNAs during messenger RNA maturation in the nucleus. Moreover, *PABPN1* is involved in regulating poly(A) tail length and mRNA stability, as well as in mediating mRNA transport from the nucleus to the cytoplasm and degradation [24]. OPMD is a rare, late-onset autosomal dominant neuromuscular disorder characterized by proximal muscle weakness, ptosis, and difficulty in swallowing [25]. The molecular mechanism of this pathology is associated with an abnormal expansion of the GCN trinucleotide repeat (between 11 and 18 repeats) in the *PABPN1* gene [26]. The pathophysiology of OPMD involves repeat expansions in the *PABPN1* gene, resulting in the production of a mutated protein that forms toxic aggregates in muscles, disrupting RNA processing and causing progressive muscle degeneration [26].

Despite being distinct disorders, OPMD and Pompe disease present significant clinical overlap [1]. These patients share clinical features such as progressive muscle weakness, dysphagia, and, to a lesser extent, ptosis and proximal muscle involvement. However, Pompe disease is distinguished by a more significant respiratory engagement and a more generalized muscle weakness [1,27]. Overall, Case 2 illustrates the importance of considering a differential diagnosis when faced with ambiguous enzymatic and genetic results.

On the one hand, this case allowed us to confirm that the patient’s decreased *in vitro* acid alpha-glucosidase enzymatic activity was due to the pseudodeficiency variant in homozygosity and that it was not associated with the patient’s final diagnosis which was OPMD [10]. Hence, caution should be taken when considering these cases, as pseudodeficiency alleles can mask the underlying cause of the disease if they are not carefully evaluated using 4-MUG as a substrate [10]. Moreover, if the patient in case 2 had been diagnosed with LOPD, she would have undergone unnecessary enzyme replacement therapy, while the OPMD would have remained undetected. On the other hand, patient 2 serves as an example exome analysis scope, as it permits *in silico* panel customization to find alterations in genes that are not the primary clinical suspicion.

In summary, we have established a global consensus on the classification of the NM_000152.5(*GAA*):c.271G>A, p.Asp91Asn variant as benign considering all the available evidence to date, analyzed in accordance with the latest international guidelines. The experience gained reaffirms that the accurate diagnosis of rare genetic diseases requires adherence to international consensus guidelines and a thorough evaluation of genetic variants. This helps prevent diagnostic errors and improves the quality of medical care. Furthermore, precise genetic diagnosis and strict variant curation are crucial for guiding therapeutic decisions and avoiding inappropriate treatments that may arise from incorrect diagnoses. Ensuring that variants are accurately classified allows for better-informed clinical choices, ultimately benefiting patient outcomes by tailoring therapy to the correct underlying condition. Finally, sharing data with reliable results is vital for future reinterpretations of variants, so we encourage the scientific and medical community to contribute to this mission.

## Data Availability

All data produced in the present study are available upon reasonable request to the authors

https://www.ncbi.nlm.nih.gov/clinvar/variation/4020/

https://databases.lovd.nl/shared/individuals/00453583

## Data availability statement

The genetic variant data that support the findings of this study are openly available in LOVD3 database: *GAA* and *PABPN1* variants submission link: https://databases.lovd.nl/shared/individuals/00453583.

## Acknowledgments

We would like to thank the Lysosomal Diseases Expert Panel (ClinGen LSD VCEP) [https://clinicalgenome.org/affiliation/50009/] for the recommendations on classifying the NM_000152.5(*GAA*):c.271G>A, p.Asp91Asn variant. We thank Dr. Eleonora Katz for reviewing and editing the English writing of this work.

## Author Contribution Statement

Conceived and designed the methodology: F.G. and P.I.B. Analysis of clinical history and medical aspects: F.G., G.C. and C.L.M. Analyzed patient samples: F.G. and M.C. Analyzed and discussed the data: F.G., P.I.B., G.C., C.L.M., V.D., L.L. and M.C. Wrote the paper: F.G., P.I.B., V.D. and M.C. All the authors reviewed the manuscript.

## Ethical Approval

The study was conducted in accordance with the Declaration of Helsinki and approved by the Institutional Review Board (or Ethics Committee) of Facultad de Farmacia y Bioquímica (06122023-167) and by the Institutional Review Board (or Ethics Committee) of Hospital de Clínicas “José de San Martín” (1218-23).

